# Exploring athlete pain assessment experiences and priorities; A two-part qualitative series of athlete and physiotherapist interactions. Part Two. “Forging Our Future” - Athlete and physiotherapists’ priorities for pain assessment and beyond

**DOI:** 10.1101/2024.01.20.24301522

**Authors:** Ciarán Purcell, Caoimhe Barry Walsh, Garett Van Oirschot, Brona M Fullen, Tomás Ward, Brian M Caulfield

## Abstract

**Objectives:** To explore the priorities and directions of athlete upper and lower limb pain assessment by facilitating shared understandings of athletes and sports physiotherapists.

**Design:** Qualitative Research using a hermeneutic phenomenological approach.

**Methods:** We carried out focus groups using a deliberate criterion sample and a constructivist perspective. At the end of each focus group, we used the nominal group technique method to generate a list of consensus-based priorities for future pain assessment. Our paper follows the consolidated criteria for reporting qualitative research (COREQ) guidelines.

**Results:** We completed five focus groups, comprising twelve athletes (female, n=5, male n=7) and four sports physiotherapists (male, n=4) Two final themes (and six subthemes) were developed; (i) Enhanced Communication and Pain Descriptions (describing and representing pain, better communication, the role of technology, providing direction and setting the pace), (ii) Integrating Sport Specific and Multidimensional Assessments (broadening the pain assessment toolkit, the role of technology). We developed a set of thirteen practical priorities for pain assessment that span the subjective, objective, and general aspects of the athlete pain assessment.

**Conclusion:** We have presented stakeholder-generated perspectives, directions, and priorities for athlete pain assessment. Athletes and Physiotherapists must continue to work together to achieve a comprehensive sport-specific multidimensional pain assessment experience alongside their wider support networks to ensure optimal representation and communication. We have highlighted some available pain assessment tools and strategies and outlined how novel tools may address certain gaps in the assessment process. Researchers, clinicians, and athletes can consider the practical guidance we have provided to address these priorities.

## Introduction

In this series, we acknowledge the value of qualitative research in sport, and we are pursuing a comprehensive understanding of athlete pain experience and assessment through qualitative methods.^1^ The International Association for the Study of Pain (IASP) outlines the importance of appreciating and validating each individual’s experience, including both the sensory and emotional aspects of pain in their definition of pain.^2^ The contemporary predictive processing model of pain describes how pain is not merely something we experience but something that influences how we act and interact in the world, shaping our behaviours.^3^ Pain neuroscience has been applied to athlete cohorts with guidelines recommending a multidimensional biopsychosocial approach to facilitate optimal pain assessment.^4–6^ Contextual pain assessment tools ( affective, cognitive and socioenvironmental) have been used substantially less frequently than the more traditional tools (neurophysiological and biomechanical pain assessment) in both research and practice settings over the past fifty years and, somewhat concerningly, the gap between the use of these wider aspects and the more commonly used traditional tools is widening.^7^ In Part One of this series, athletes and physiotherapists shared their experiences of the content and qualities of pain assessments. Athletes and physiotherapists discussed the commonly used tools, measures and scales highlighting the strengths and limitations of current practice in the context of best available guidance. The quality of the pain assessment was closely linked with the pain interview, an opportunity for athletes to tell their stories, share information related to the affective, cognitive, and socioenvironmental aspects of pain and develop a strong therapeutic alliance. These findings align with recent literature emphasising the power of narrative methods to express the complexity of feelings, emotions and experiences influenced by sport-specific sociocultural aspects that athletes negotiate daily. ^8–10^ In this paper, the second in this two-part series we present the priorities for future practice as part of an integrated overview and culmination of our overall findings.

## Methods

### Focus Groups

We carried out focus groups with a deliberate criterion sample of athletes and sports physiotherapists based in Ireland from diverse sporting backgrounds. We developed a topic guide which guided discussion from broad pain experience-related questions to more focused questions on priorities for pain assessment. A moderator and neutral observer helped to ensure equity of participation and full exploration of the topic guide. We used reflexive thematic analysis and developed codes, candidate themes and final themes in an iterative fashion.^11^ A critical friend (CBW) independently reviewed the data and added additional perspectives. We have described the methodology for the focus groups component of this paper in detail in Part One of this series.^12^ In the curent paper, Part Two of the series, we use data from focus group questions that address priorities and directions for athlete pain assessment. The full published data set can be accessed at https://data.mendeley.com/datasets/t47tw94mzd/2.).

### Nominal Group Technique

Nominal Group Technique (NGT) is a method of problem-solving and idea generation where all participants have an equal opportunity to contribute their priorities, effectively minimising power imbalances and facilitating the ranking of priorities suggested by participants.^13^ We used the NGT at the end of each focus group session to facilitate brainstorming of priorities and generate consensus for items to be included in the next phase of an athlete upper and lower pain assessment framework development. We chose this method to give equal weight to the opinions of all participants and to focus the ideas and topics discussed in the focus groups on priorities for pain assessment practice. NGT is a method of consensus generation where all participants contribute multiple ideas which are then voted on and prioritised equitably.^14^ Members rank their top five with their highest priority idea receiving five votes down to their fifth highest priority idea receiving one vote. NGT consists of four stages: Silent generation, round robin, clarification, and voting. We used five sequential steps (state the subject, reflection and writing, polling, discussion, and prioritisation) to operationalise this technique which was first developed in 1975 and has been used across domains and settings including healthcare and sport.^15,16^ We recorded and discussed all ideas generated by participants during the polling stage of the NGT. Participants voted on initial ideas which then became priorities. We discarded all ideas that did not receive a minimum of one vote during this voting stage from our analysis. See Appendix A for a detailed description of how we applied each of the five steps in this study. We carried out the five NGTs independently and pooled the results during the data analysis stage.

## Data Analysis

During the Focus Group data analysis process, we assigned athletes alphanumeric participant IDs beginning with the letter “A” and physiotherapists alphanumeric IDs beginning with ‘’P”. Athletes and physiotherapists were talking about and experiencing the same concepts so we merged the data for coding and analysis. Participant IDs were known only to CP, the lead researcher. Once participant IDs were allocated and analysis was complete all records of participant information were deleted ensuring anonymity.

After we completed the NGTs, we reviewed all priorities that received a minimum of one vote, and I (CP) applied the initial codes. We (CP, BC, TW and GvO) reviewed the initial coding framework. We updated the codes and renamed them where necessary, grouping similar codes to generate candidate themes of assessment items and aspects. CBW reviewed the codes and candidate themes independently in her role as a critical friend and we discussed the additional perspectives. We then updated our candidate themes and developed a set of finalised themes.

## Results & Discussion

The results are derived from five focus groups which gathered the experiences and interactions of sixteen participants (athletes, n=12, physiotherapists, n=4) from a broad range of sports and competition levels, a full description of which is provided in Part One of this series.^12^ Athletes and physiotherapists voiced their opinions on what the future of an athlete pain assessment could and should look like. From the focus group codes, we developed two clear themes highlighting priority areas for pain assessment.

In Figure 1 we present these two themes and their associated codes. The first theme centres around better strategies to describe and represent pain, including tools that go beyond the current methods for capturing pain intensity and enhanced pain communication strategies. Participants highlighted aspects such as the context, timing and frequency of pain assessments and proposed technology-based solutions as one option to address current shortcomings. The challenge and opportunity of providing athletes with clear direction in the next steps of their pain assessment and management process are highlighted. The second theme explores prioritising assessment strategies for the different aspects of pain experience. Future pain assessments should be multimodal, considering a wide range of biopsychosocial and sports-specific factors to get a more comprehensive picture of the pain the athlete is experiencing. Both strategies promise to enhance current assessments offering a deeper and more nuanced understanding to better guide decision-making and management.

**Figure 1.**
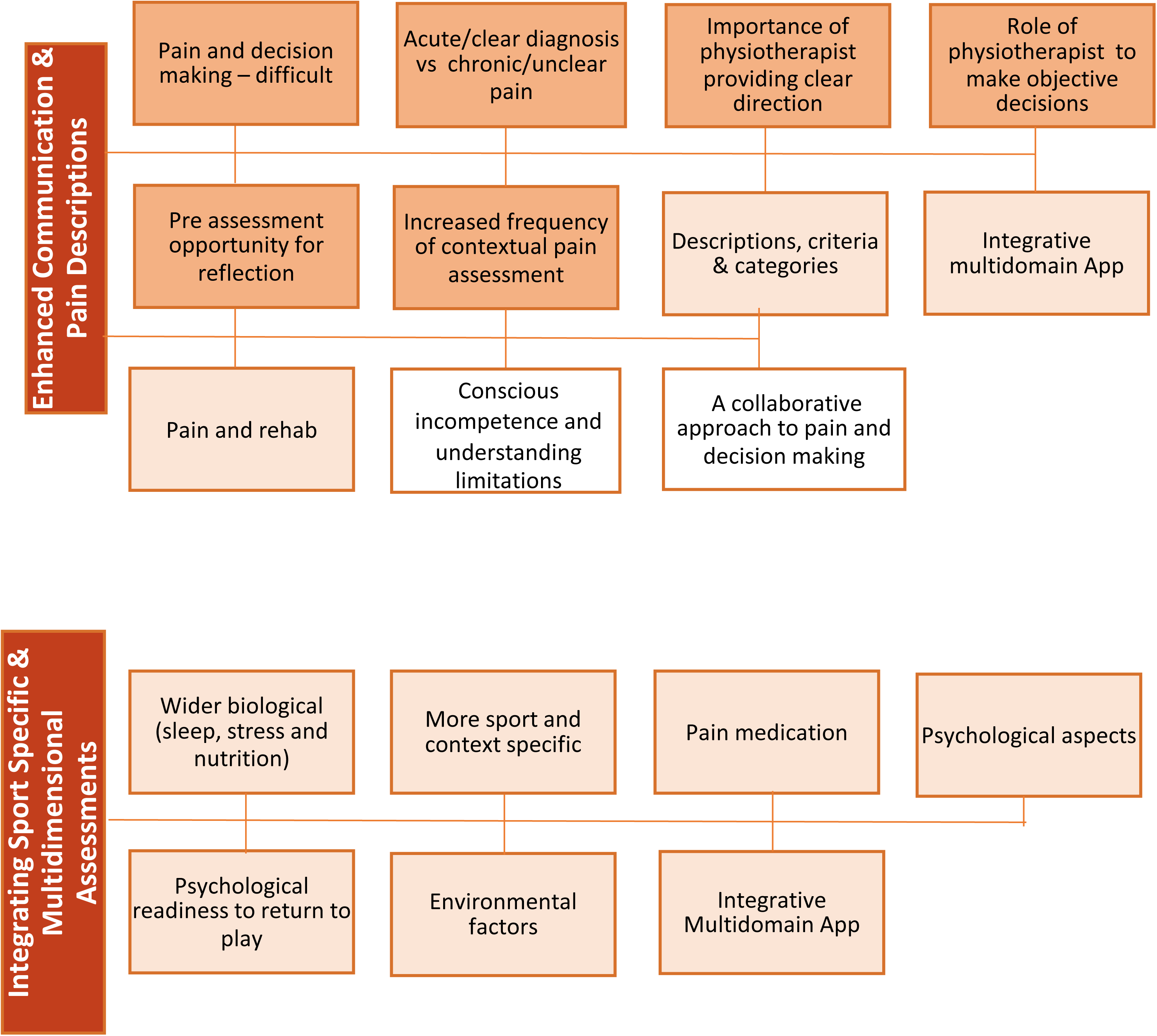
Themes (Enhanced Communication and Pain Descriptions; Integrating Sport-specific and Multidimensional Assessments) and codes. Dark shading – indicates codes that were present in athletes and physiotherapists. Light shading – indicates codes that were present in athletes only. No shading – indicates codes that were present in physiotherapists only

In Figure 2 we present the finalised thematic map of the athlete pain assessment priority pyramid which demonstrates how the themes and subthemes explored in this paper build on Part One of the series.

**Figure 2.**
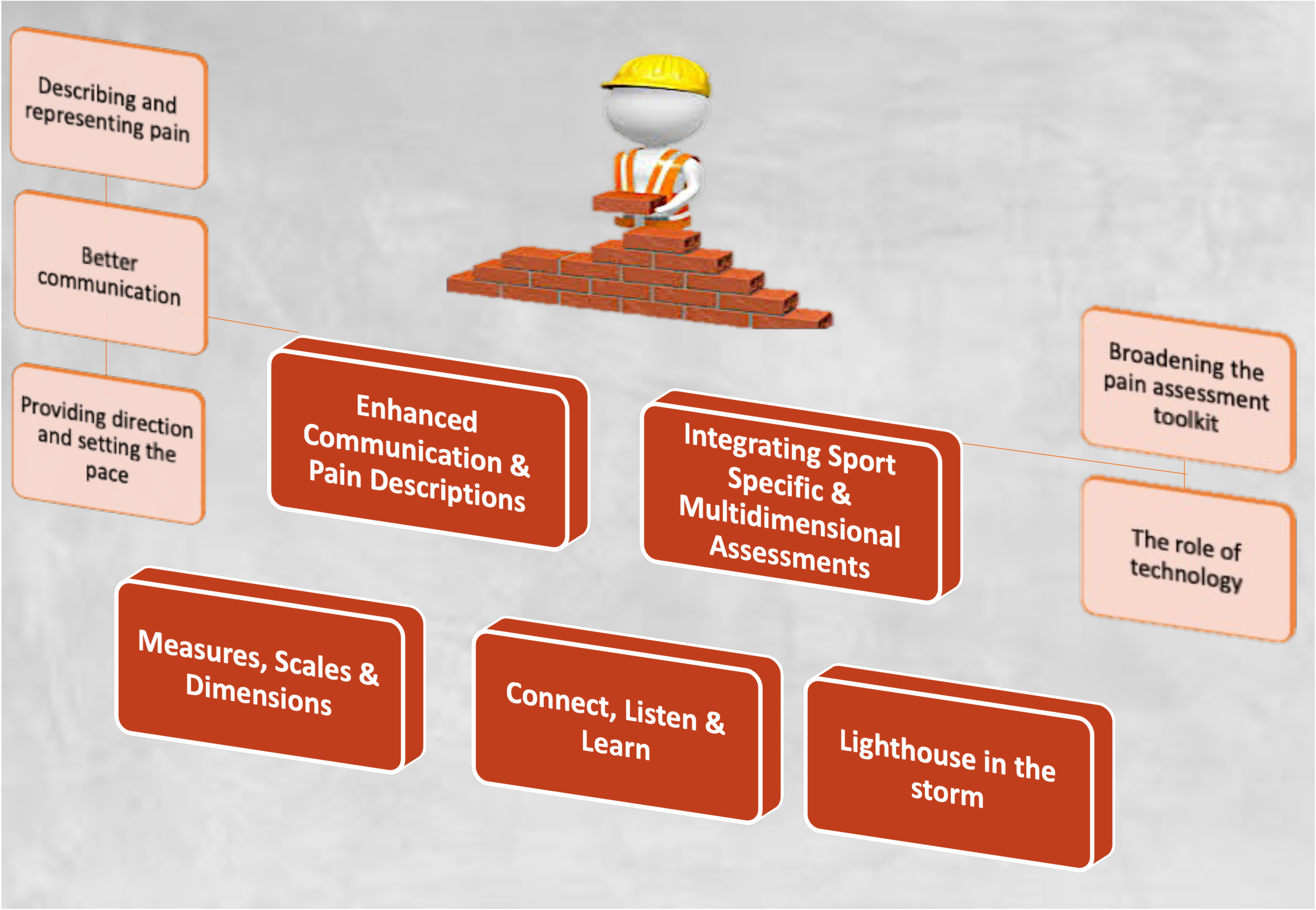
Athlete pain assessment priorities themes and subthemes. The themes for each part of this series represent a row in the priorities for pain assessment pyramid, this paper presents the top row of the pyramid, the themes and subthemes that address the priorities for athlete pain assessment. These themes build on the bottom row or foundation of the pyramid presented in Part One. Source: The cartoon element at the top of this image was designed by Freepik www.freepik.com

### Theme 1 Enhanced communication and pain descriptions

Theme 1 includes three sub themes; 1.1 – describing and representing pain, 1.2 – better communication and 1.3 – the role of technology.

#### 1.1 Describing and representing pain

Participants discussed the struggle to describe and represent pain, and how it may be improved by using more comprehensive descriptions. This reflects the contemporary drive for valuing the patient’s language and selecting appropriate metaphors to help represent and explain the pain experience.^17^ Additionally athletes felt providing contextual information such as their pain and rehabilitation history helped explain their current pain experience. Athletes and physiotherapists highlighted the development of criteria and categories for the classification of pain intensity that go beyond the traditional numerical pain rating scale (NPRS) to include more detailed descriptions of pain and the impact of pain on aspects of daily life as a priority for pain assessment.

> “I think more kinds of adjectives describing pain it definitely would be beneficial.” - A09

> “Your rehab experience. Because I think they all play into how you experience pain and maybe your history of pain or your even perception of pain because everyone is an individual and they experience lots of things differently.” - A02

> “It would be great to see something where it was like how does this pain influence and then there’s like several categories so maybe one would be sleep for me anyway, one would be like work you know or your sport…” – A10

There may be value in revisiting long-established measures such as the Brief Pain Inventory Scale (which includes intensity, location, body chart, interference with everyday life and pain within a certain timeframe) and the McGill Pain Questionnaire (which includes a comprehensive list to describe pain sensation, physical and emotional aspects, interaction with treatment options and wider lifestyle factors and measures of intensity that provide context to this pain experience) and developing contemporary multidimensional tools tailored to an athlete population.^18,19^

#### 1.2 Better communication

The timing, setting and communication of pain were earmarked by participants as areas to improve athlete pain assessments and enable some of the proposed enhanced descriptions and categorisation. Athletes and physiotherapists felt including more time for reflection, both before and after the athlete-physiotherapist assessment session, would facilitate athletes to recall their pain experiences in greater detail. Additionally, varying the time of assessments to capture pain during competition and activities of daily living that provoke pain was suggested as an opportunity to enhance pain assessment practice, particularly when athlete-physiotherapist interactions are infrequent, and athletes may struggle to accurately recall the entirety of their pain experience since their previous assessment.

> “I like the idea of writing stuff down before you go in and see someone because you get a chance to actually think of it.” – P04

> “I know that helped for me, because.. before I go into a physio, I do write down what I’m going to say, because more often than not I’d leave forgetting to mention things and you’re like, oh god, well it’s gone now and I’m never going to say anything.” - A11

Ecological momentary assessment (EMA) involves the repeated sampling of behaviours and experiences in real-time and is carried out in the person’s natural environment.^20^ EMA has been applied in chronic pain and sports psychology research and our results suggest it is something which may provide value when considering improved communication in future pain assessments for athletes. ^21,22^ Participants felt applying enhanced communication methods would help with difficult decision-making when it comes to athlete pain. Athletes described the challenge of not knowing when to play or compete through pain and how to effectively gauge progress whilst physiotherapists noted the opportunity for additional communication to enhance this process.

> “I think as an athlete .. we want to know .. when is that pain too much to keep doing what I’m doing, when should I stop? That type of thing like what’s the recovery process?” – A06

> “Sometimes as physios that’s where we could maybe have that missing link…, not necessarily to be so individualised and so 24/7 care but there has to be some level of a way of you know being able to kind of link in with them I think a bit more that I think would help sometimes.” - P02

#### 1.3 The role of technology

The effective use of technology to facilitate enhanced communication and contextual assessment was proposed as a future opportunity. Technology could be used to integrate novel assessment methods such as EMA. Athletes based in high-performance environments discussed how they used smartphones to facilitate more frequent communication about pain with their sports medicine teams which they found helpful. The value of insights gleaned from regular subjective wellness reporting with athletes has been established and has been proposed to provide more value than objective measures for certain training load, health, and well-being markers.^22^ However, further consideration is needed to smoothly integrate technology into the contemporary pain assessment process and wider athlete ecosystem. Aspects such as validity, data protection and integration into current workflows and health system records all warrant consideration. The clinical utility of a single self-report item has been challenged in line with the need for interpretation and reasoning when it comes to musculoskeletal clinical practice standards.^23,24^ Furthermore, the future integration of technology must meet the needs of both athletes and physiotherapists, enhancing rather than complicating the pain assessment clinical encounter.

> “An app we use for training, and she (coach) would put in each day like how many hours did you sleep last night? What’s your perceived rate of recovery from the previous day’s training? Perceived rate of like pain that day.” – A08

> “Could you fill in maybe an assessment after each training session, describing your pain and send that back to the physio or have regular updates throughout that process as well?” – A05

> “A pain app where every morning it was what’s your pain and then a journaling section and it’s like how did you sleep last night? Did you take painkillers? How was your training? All these different sections so and then say next time you see your physio potentially they would have access to your app and it’s like okay here’s with your rating over the last week” – A10

#### 1.4 Providing direction and setting the pace

An effective assessment sets the pace and expectations for the management process. Athletes found that receiving an accurate prognosis and timeline to build goals as a pivotal part of pain management. Athletes and Physiotherapists highlighted that the role of the physiotherapist is to provide an objective perspective to aid in the decision-making process. The optimal level of objectivity and guidance varied amongst athletes and sports environments. Regarding priorities for pain assessment practice, physiotherapists acknowledged the importance of providing clear direction at the end of the assessment.

> “I think clarity is probably the most important thing, just having a timeline said to you that you can believe and actually see is realistic, I think that’s very important in terms of progressing” – A05

> “As much as you’re a fan of the sport and a runner as well you kind of have to then be able to stand aside and go hold on what’s the best for the athlete” – P01

> “You’re having issues with these three areas, this is what we’re going to work on..” – P04

However, both athletes and physiotherapists emphasised how diagnoses, timelines and decision-making are not always straightforward. Physiotherapists found making a definitive diagnosis and providing direction was particularly challenging in persistent pain presentations, where the signs and symptoms that frequently accompany acute pain and injury presentations were not present and recovery and improvement were often protracted requiring a greater level of pain knowledge and clinical reasoning skills. These challenges align with gaps and future priorities in athlete pain assessment and management identified by the International Olympic Committee (IOC).^25^

> “I agree completely with what P03 was saying as regards how cloudy things become over time and I think that that’s a difficulty like for … with a patient… I would always say to them .. look the longer we go away from your injury to where we are now, the more likely is we won’t be able to give you a definite diagnosis of what’s going on” – P02

Acknowledging these challenges in an area that does not always have a clear answer, physiotherapists prioritised understanding their limitations as an important aspect of the role of the sports physiotherapist. Additionally, they highlighted the importance of honesty and focusing on aspects within their remit to facilitate the development of the trust required for an effective athlete-physiotherapist assessment and relationship.^26^

> “Lads come in with pain and ask what is it?..I don’t know exactly what it is, it could be this, this and this, but all I know is that you’re having issues with these three areas, this is what we’re going to work on..we’ll see what it’s like.” – P04

> “It’s okay to not know and I think sometimes it’s better to tell someone you don’t know but definitely have a direction you feel like they need to go in” – P04

### Theme 2 Integrating sport-specific and multidimensional assessments

Theme 2 includes two subthemes: 2.1 – broadening the pain assessment toolkit and 2.2 – the role of technology. In parallel to enhanced communication and descriptions, athletes felt multimodal and sport-specific pain assessment tools would give a more comprehensive assessment and understanding of the athlete’s pain experience. This theme was developed from codes gleaned from athlete perspectives only and so it is a truly athlete-focused theme.

#### 2.1 Broadening the pain assessment toolkit

Psychosocial, lifestyle and environmental pain assessment tools were recommended by athletes to give a more encompassing and holistic overview of athlete pain. Athletes highlighted sport-specific pain assessment tools such as assessing movement, strength and motor control patterns relevant to their sport, asking about training load and the impact of pain on sports activities and measuring pain during, before or after sports activities to represent the unique athlete pain experience and provide enhanced contextual information in line with the best available athlete pain guidance.^4,27^ Athletes discussed how clinic-based pain assessments could be enhanced by completing more assessments in the athlete’s sports environment such as on the pitch, court or track, a finding that could facilitate the practical implementation of contemporary IOC guidance.^4,27^

> “The other environmental or the psychological factors that could have led to it because sometimes it’s a very obvious thing, that reason it happened. But asking about how you feel around it and a little bit of support…getting a good grasp of someone’s life and all the other stressors and factors I think definitely would help in diagnosing the thing rather than you know just going for more scans and what have you to try to pinpoint it.” – A09

> “The questions that they use for pain, there’s no differentiation between some guy who hurt his leg working on a building site versus somebody who’s like an amateur athlete who’s you know playing sport five nights a week and the questions need to be more specific to that person, that kind of activity maybe.” - A02

> “More assessments directed at for the readiness to play in terms of I think even out on a pitch is completely different in the physio room .. when you’re out on the pitch afterwards and you feel different types of pain .. could the physio be out and involved more?’’ - A05

#### 2.3 The role of technology

Technology was once again suggested by athletes as a potential method to add multimodal and sport-specific measures into an integrative pain assessment. Additionally, the role of the physiotherapist and their expertise in pain assessment and management was stressed, and future pain assessment strategies should augment rather than replace this vital relationship. Additionally, a recent systematic review found that there is moderate to high certainty evidence supporting the integration of technology into a physiotherapy assessment for musculoskeletal disorders in the general population.^28^ The study found substantial to excellent validity and excellent inter and intra-rater reliability for pain (97-98% agreement) and patient-reported outcome measures (ICC 0.99-1.00, 95% CI 0.97 to 1.00). However, patients reported a superior user experience and confidence in the examination from a face-to-face assessment, supporting our findings. Additionally conducting and interpreting pain assessments with athletes requires sports and context-specific information (such as sport-specific physical demands, and coach and team expectations for return to play) that may be understood better if conducted in the athlete’s sporting environment.

> “So, if I could see changes in the industry its App based for sure it’s probably interactive ..but there’s some clear definitive I want him (the physiotherapist) to have a relationship with me, know what I’m doing and know when I can get back to my sport so there’s something in that. I don’t know what but…” – A06

### From theory to practice: practical priorities for pain assessment

In Table 1 we present the pooled results of the nominal group techniques we carried out. Each theme is a practical example of an aspect of assessment that can be used to operationalise the pain assessment priorities we identified through the focus group findings above. We present the codes that comprise each theme alongside a qualitative description of how clinicians can apply the assessment item in practice. We have indicated whether each item can be incorporated as part of the subjective assessment (initial interview), objective assessment (physical testing) or should be considered as a general aspect of the wider assessment process. The proportion of total votes each theme received when codes were pooled is also presented. The majority of the practical pain assessment priorities we present align closely with the published literature including 1) subjective components such as; establishing the history, context, characteristics, severity and impact of the athlete’s pain and exploring the athlete’s stressors, psychological aspects and lifestyle factors as well as their training load and 2) objective components such as; assessing and identifying pain through movement, completing sports specific objective measures both at the site of pain and throughout the kinetic chain. ^2,4,25^ Developing a specific, clear, and time-appropriate assessment is a novel finding. Although the IOC guidelines recommend tailoring the pain assessment based on the presentation of the athlete and the stage in the pain management process our findings add that the assessment should align closely with the athlete’s goals and the plan should be articulated clearly to the athlete so that they understand each aspect of the pain assessment and why it is being completed. Another novel finding is the need for alternative pain severity scales that go beyond the traditional numerical pain rating scale. Scales such as the traffic light pain scale which offers athletes guidance on decision-making regarding playing through pain have clinical utility and are used widely in practice. ^29^ Additionally there is scope for the inclusion of a pain severity scale that helps to overcome the limitations of a numerical scale to describe and represent the pain experience more appropriately although what that might look like is yet unclear. The International Association for the Study of Pain (IASP) identifies how a verbal description is just one method of representing pain.^2^ Additional options include athletes using a body chart to indicate their pain or the demonstration of their pain through a specific movement or action to have their experience of pain understood and validated which is another novel finding.

**Table 1 –.**
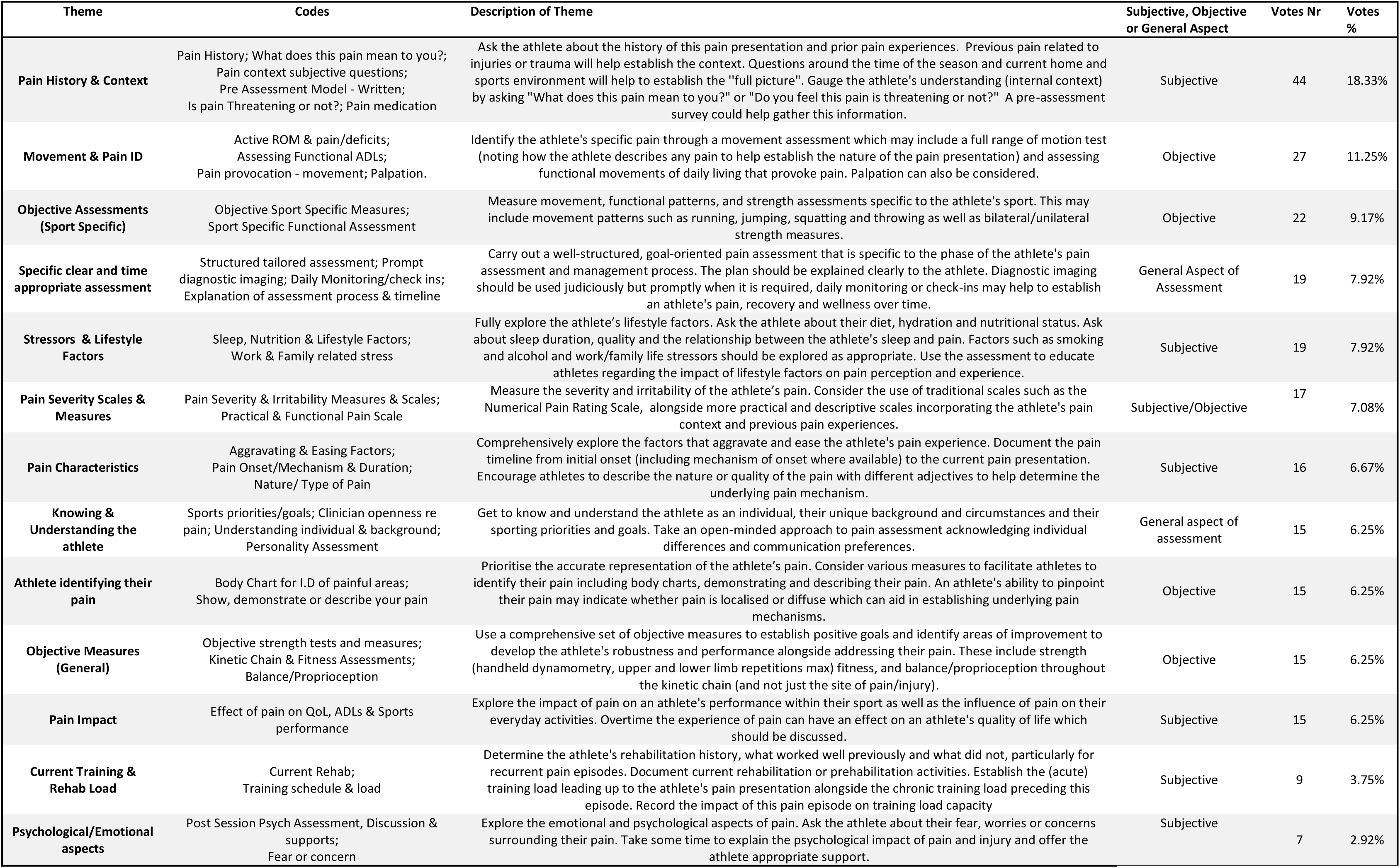
Nominal Group Technique Results inserted here. Votes Nr – is the number of votes each theme received. Each participant received a total of 15 votes which they used to rank their top five priorities/codes (5 votes, 4 votes, 3 votes, 2 votes and 1 vote.) Votes % displays the overall percentage of votes each theme received. Pain ID - Pain identification, ROM - range of motion, ADLs – activities of daily living, Qol – Quality of life.

## Conclusion

In this paper we have presented themes focused on future pain assessment priorities and directions that build on our “athlete pain assessment” findings presented in Part One of this series. Moving beyond unidimensional, point-in-time clinical pain measures to encompass better communication and direction for pain management will involve the use of more comprehensive and descriptive pain scales and tools that athletes and physiotherapists can relate to. We have outlined examples of pain assessment tools that are currently available and should be considered by clinicians. Conversely, we have highlighted how novel tools may encompass capturing pain at critical time points in sports environments to help tell the full story of athlete pain. Whilst Physiotherapists may be justifiably reticent to introduce additional measures to avoid complicating the assessment process for athletes, athletes are embracing existing and emerging technology as part of evolving sports science. Applying a carefully developed and well-implemented solution that incorporates available technology and keeps the athlete-physiotherapist interaction and relationship at its core is one potential future solution that must be considered.

Although we collected experiences from a diverse range of athletes and physiotherapists and variety was achieved in sport, competition level and practice setting the experiences and priorities gathered from these focus groups and nominal group techniques may not apply to all athlete pain assessment settings. Notably, participants were all recruited from Ireland and whilst some of the female athletes also had a physiotherapy background, no female sports physiotherapists were available to participate. The focus of our research was to highlight the athlete and physiotherapist voices and explore their experiences of pain assessment through open discussion. In addition, to augment these findings, we selected the Nominal Group Technique to complement the exploratory nature of the focus group methodology. We have presented a comprehensive series of pain assessment priorities for physiotherapists to consider. There is scope for future research to consider these findings in light of contemporary research and practice and develop an expert sports physiotherapist consensus-based athlete pain assessment framework that can be implemented in research and practice settings.

## Practical Implications

- The pain assessment must include assessment tools and measures that allow athletes to, represent and therefore validate their specific pain experience.
- Physiotherapists should use pain assessment tools that capture the context of an athlete’s pain as well as the intensity and impact on sports and activities of everyday life at key points in an athlete’s day.
- Physiotherapists must consider a variety of pain assessment tools that address the multidimensional nature of pain and integrate psychological, social, environmental and sport-specific aspects of the athlete’s pain experience in the pursuit of providing direction for athletes to manage their pain.
- Physiotherapists should consider the careful and effective use of technology to augment the athlete-physiotherapist pain assessment interaction.

## Data Availability

The full published data set can be accessed at

https://data.mendeley.com/datasets/t47tw94mzd/2.

## Appendix A – The Five Steps of the Nominal Group Technique

1. **State the subject**. The lead researcher wrote the title of the subject at the top of a whiteboard (virtual whiteboard for Zoom session) “List all the items you think should be included/prioritised in an athlete pain assessment framework.”
2. **Reflection & writing**. Ten minutes were given for silent reflection and consideration and participants were asked to write down all of their ideas on separate sticky note pages to keep their ideas private until the polling stage.
3. **Polling**. Each group member revealed one idea at a time, taking turns until every idea was recorded by placing the sticky notes on the whiteboard. Minimal discussion took place at this stage with equal contribution from each member.
4. **Discussion**. Each idea was then clarified and explained by the participant who proposed it, followed by discussion and queries by the group. The specific wording of ideas was changed in some circumstances following discussion upon approval from the person who came up with the idea. Similar ideas were merged or grouped together at this stage.
5. **Prioritisation** Following discussion each participant ranked their top five assessment priority ideas from all of the ideas generated and approved in earlier stages. A score of five was allocated to the idea the participant ranked highest, a score of 4 was allocated to the idea the participant ranked next highest and so on continuing to one. The ideas were then ranked based on the highest to lowest scoring. Participants were then invited to share any element they felt was missing or needed to be adjusted before the final ranking was approved. All ideas that received a minimum score of one vote were kept.

## Notes

### Competing Interest Statement

The authors have declared no competing interest.

### Funding Statement

This work was supported by funding from Science Foundation Ireland under the grant for the Insight SFI Research Centre for Data Analytics (SFI/12/RC/2289_P2) Funders had no role in the data collection, analysis or interpretation and will have no role in approving the final manuscript.

### Author Declarations

Permission was granted for this study by the University College Dublin Human Research Ethics Committee. (LS-22-40-Purcell-Caulfield)

### Summary of Updates

The original version was changed significantly following feedback from editors, influencing data analysis and how it is presented in results and discussion. The methods and results content remains the same.

